# Cost Minimization Analysis of Digital-first Healthcare Pathways in Primary Care

**DOI:** 10.1101/2025.04.04.25324363

**Authors:** Alexandra Dahlberg, Sakari Jukarainen, Taavi Kaartinen, Petja Orre

## Abstract

A retrospective, registry-based cost-minimization analysis assessed whether initiating acute primary care episodes via a digital-first pathway reduces costs compared to traditional care in a Finnish setting (Harjun Terveys). Of 637,923 encounters, 64,969 eligible acute episodes were identified. After propensity score matching (19,697 pairs), mean episode costs were significantly lower in the digital pathway (€170.74) than in traditional care (€220.91), reflecting a 22.7% reduction (*P*<.001). Savings varied by clinical presentation, from 10.3% for respiratory infections to 52.5% for gastroenteritis (all *P*<.001). Digital care was associated with lower use of laboratory tests and imaging. Follow-up visits were generally fewer in the digital group, except for respiratory infections, which showed a slight increase. Sensitivity analyses with 7- and 30-day follow-up windows confirmed the findings. Overall, this study supports digital-first models as a cost-effective strategy for managing acute conditions in primary care, with potential to reduce unnecessary resource use without compromising continuity of care.

## Introduction

Healthcare systems globally face growing financial pressures due to aging populations^1^, increasing prevalence of chronic diseases, and escalating healthcare costs. These pressures underscore an urgent need for innovations that enhance efficiency and cost-effectiveness in care delivery. Digital health technologies—including telemedicine, mobile health applications, and online patient portals—have emerged as promising solutions to deliver primary care more efficiently^2–4^. By reducing overhead, optimizing provider time, and streamlining patient management, digital interventions have the potential to decrease overall healthcare expenditures. Systematic reviews have indicated that many digital health interventions can be cost-effective and sometimes even cost-saving^5,6^. However, real-world evidence demonstrating economic benefits in routine healthcare settings remains limited, highlighting the need for rigorous economic evaluations in practice^7^.

One analytical approach for comparing costs in such settings is cost-minimization analysis, which is appropriate when clinical outcomes of the interventions are expected to be equivalent^8^. In the context of acute, minor health issues (for example, managing an uncomplicated infection via a digital visit versus an office visit), the health outcomes are generally assumed to be comparable. Thus, any difference in cost between a digital and a traditional pathway would primarily reflect differences in efficiency rather than effectiveness. Focusing on cases where outcomes are equivalent allows us to directly evaluate whether digital delivery truly economizes resources compared to traditional care. However, confounding factors may influence the analysis when comparing digital and traditional care, as patients choosing digital pathways may differ in demographic or clinical characteristics^9–11^. To address this, propensity score matching (PSM) was applied to account for these factors.

Building on this analytical framework, this study addresses a critical gap by examining the cost-efficiency of digital healthcare pathways implemented in a large-scale primary care setting in Finland. In the Päijät-Häme Wellbeing Services County in Finland, patients with acute primary care needs can seek care either by using a digital clinic app (Päijät-Sote) or through traditional means like calling a health center or walking in. Leveraging this parallel delivery system, the study evaluates whether initiating care digitally results in lower total episode costs compared to traditional methods for common groups of acute conditions, such as dermatological symptoms, gastroenteritis, ophthalmologic symptoms, respiratory infections and urinary tract infections. This was achieved analyzing real-world cost and utilization data from a Finnish case study.

## M ethods

### Study Design and Setting

We conducted a retrospective registry-based study with a cost-minimization analysis framework^12^. This approach was selected based on the premise that both digital and traditional care pathways manage the same acute conditions under standardized clinical guidelines, with the expected outcome being patient recovery. Given this assumption of equivalent clinical effectiveness, cost-minimization analysis allows for a direct evaluation of economic efficiency by comparing healthcare expenditures between the two pathways.

The study was conducted in the Päijät-Häme region of Finland, within Harjun terveys a public-private joint venture, which serves the municipalities of Lahti, Iitti, Kärkölä, and Hartola, covering a population of approximately 134,000. Harjun terveys provides typical Finnish publicly funded primary healthcare services and has increasingly adopted digital services in recent years. A digital primary care platform (the Päijät-Sote digital clinic app) was introduced in this region on May 1, 2023, operating alongside traditional care access. The study observation period ran from May 1, 2023 through September 30, 2024, covering the first 17 months of the digital clinic’s implementation.

This research followed the Consolidated Health Economic Evaluation Reporting Standards (CHEERS) guidelines^13^. The study was approved by Päijät-Häme Wellbeing Services County. Individual patient consent or separate ethical committee approval was not required under the Finnish Act on the Secondary Use of Health and Social Data (552/2019)^14^. All data use complied with relevant data protection regulations (including the EU GDPR)^15^.

### Inclusion Criteria

All patients with their home address in the catchment area and who were alive for study period were included. All primary care contacts with nurses and doctors were included, except contacts related to health promotion (e.g., vaccinations, maternity and child health clinics). Secondary, tertiary, and inpatient care were not analyzed. We defined an “episode” as a fixed period of 14 days following the initial primary care contact. We restricted the analysis to five categories of clinical presentation: dermatological symptoms, gastroenteritis, ophthalmologic symptoms, respiratory infections and urinary tract infections. These categories were chosen to represent typical acute problems commonly managed in primary care and amenable to digital consultation (e.g., no definite need for a physical examination). Episodes were included if the primary diagnosis at the index encounter matched one of the predefined categories, as identified by ICD-10 and ICPC-2 codes in the registry (Supplemental Table A1). Each episode’s care utilization was then tracked for 14 days from the initial contact. Finally, for urinary tract infection episodes, we excluded patients over 65 years old; national clinical protocol does not recommend remote management of UTIs in that age group.

### Digital vs. Traditional Pathway Definition

The digital pathway was defined as episodes where the patient initially contacts the healthcare provider through the Päijät-Sote digital platform. On the platform the patient or guardian on behalf of the patient initiates the digital contact either by filling out an automated symptom assessment or by describing one’s symptoms with free-form text. A registered nurse then performs a care-need assessment by reviewing the completed questionnaire or the free-form text and by interviewing the patient via chat when necessary. Nurses have the ability to consult a physician when needed, in cases requiring a prescription or diagnostic confirmation. If the situation warrants it, patients can also be transferred to a digital physician’s appointment, issued laboratory test orders or referred to secondary care or medical imaging – all delivered digitally within the same care continuum. The traditional pathway episodes began with either a phone call to a clinic or an in-person visit, without prior digital interaction. Importantly, after the initial contact, patients in both pathways could receive additional care as needed, including follow-up visits (digital or traditional), tests, and referrals.

An episode was established when a patient first contacted healthcare services via either the digital or traditional pathway. To ensure independence, episodes were only included if no prior healthcare encounters had occurred within the preceding 14 days. Each episode was followed for 14 days, capturing all subsequent healthcare interactions related to the initial diagnosis. Our cost analysis encompassed all primary care services utilized within this 14-day follow-up window, irrespective of modality, to fully capture total episode costs. The first ICD-10 or ICPC-2 code present during an episode was used to determine the clinical presentation-category, so for example if the first encounter had no recorded code the next encounter with a code was used.

### Cost Estim ation

We obtained cost data from the provider’s administrative database and regional cost catalogs. Each healthcare encounter in primary care has an associated cost based on standard tariffs (including staff time, facility use, etc.), and each lab test and imaging study has a unit cost. We used the Päijät-Häme Wellbeing Services County pricing catalogue for public primary care services to assign costs (Supplemental Table A2)^16^. These prices reflect the health system’s perspective (approximately the cost or tariff charged for providing the service). Costs were calculated in Euros (€) at 2024 price levels. For each episode, we summed: (1) all encounter costs (clinic visits, digital consultations, phone consultations), (2) any laboratory test costs ordered during the episode window, and (3) any imaging costs (e.g. if an X-ray was done). We did not include indirect costs (like patient travel expenses or lost productivity) in the base analysis – our perspective is that of the healthcare provider/payer.

### Outcome Measures

The primary outcome was the difference in mean total episode cost between digital and traditional pathway episodes, for each diagnostic group and overall. We calculated mean cost per episode for each group in each pathway, and the percentage difference (cost saving) of digital vs. traditional. Secondary outcomes included the breakdown of cost components (encounter vs. lab vs. imaging) by pathway, and the rate of follow-up visits in 14 days (to check if one pathway led to more revisits).

### Statistical Analysis

After forming the episodes, each patient might have zero or more episodes during the observation period. To control confounding factors, we applied 1:1 propensity score matching (PSM) to match digital and traditional episodes^17–19^. Propensity scores were estimated with a logistic regression model, using age, sex, Charlson comorbidity index^20^, and number of primary care doctors’ visits in the 2 years prior to start of the observation period (May 1, 2023)^20^. Quadratic and cubic terms were included for age and for the prior visit count in the propensity model. Age was excluded from calculating the CCI as it was included as a separate covariate. The CCI score was restricted to have a maximum of 3 due to heavy skewness.

Episodes were matched within the types of clinical presentation. Each digital episode was matched to one traditional episode with the closest propensity score (nearest-neighbor matching with caliper = 0.2)^21^. Using the matched pairs of episodes, we compared costs between pathways with independent two-sample *t*-tests^22,23^. We report on means, *t*-statistics and *P*-values for the cost differences. A *P*<.05 (two-tailed) was considered statistically significant. CCI calculations were performed using an R package ^24^, while other statistical analyses were conducted using Python software^25,26^.

## Results

### Study Population

After applying inclusion criteria and propensity score matching (PSM), the final study population consisted of 19,697 matched episode pairs, totaling 39,394 episodes. Each matched pair comprised one episode initiated digitally and one initiated through the traditional pathway, matched on patient characteristics and diagnostic category. The matched episode pairs were distributed as follows: 6,927 dermatologic symptoms, 312 gastroenteritis, 1,461 ophthalmologic symptoms, 9,616 respiratory infections, and 1,381 urinary tract infections (Table 1).

**Table 1.** Characteristics before and after PSM.

| Pathway | Unmatched |  |  | Matched |  |  |
| --- | --- | --- | --- | --- | --- | --- |
|  | Age, mean | Female (%) | N of Episodes | Age, mean | Female (%) | N of Episodes |
| Dermatological symptoms |  |  |  |  |  |  |
| Digital | 29.5 | 62.9 | 7,890 | 31.7 | 60.3 | 6,927 |
| Traditional | 56.8 | 57.1 | 19,008 | 32.7 | 60.9 | 6,927 |
| Gastroenteritis |  |  |  |  |  |  |
| Digital | 23.4 | 61.1 | 804 | 28.6 | 63.1 | 312 |
| Traditional | 37.6 | 57.0 | 393 | 28.8 | 57.4 | 312 |
| Ophthalmological symptoms |  |  |  |  |  |  |
| Digital | 24.9 | 63.9 | 3,149 | 35.0 | 62.6 | 1,461 |
| Traditional | 51.1 | 64.9 | 2,454 | 36.2 | 63.0 | 1,461 |
| Respiratory infections |  |  |  |  |  |  |
| Digital | 25.6 | 63.0 | 9,616 | 25.6 | 63.0 | 9,616 |
| Traditional | 36.8 | 60.9 | 17,958 | 25.7 | 63.0 | 9,616 |
| Urinary tract infections |  |  |  |  |  |  |
| Digital | 33.3 | 92.1 | 1,933 | 35.4 | 89.6 | 1,381 |
| Traditional | 39.8 | 84.4 | 1,764 | 35.0 | 89.1 | 1,381 |

Before matching, patients initiating care via the digital pathway were consistently younger across all diagnostic groups compared to those choosing traditional pathways. Additionally, the gender distribution varied, with the highest proportion of female patients observed in the urinary tract infection category. Post-matching analysis successfully balanced demographic characteristics between digital and traditional groups, facilitating accurate comparisons of healthcare utilization and associated costs.

Healthcare utilization metrics also showed clear differences before matching (Table 2). The traditional pathway exhibited higher mean encounters, laboratory tests, and imaging procedures in most diagnostic categories compared to the digital pathway. After matching, utilization metrics across both pathways were better aligned, allowing for direct cost comparisons attributable to the pathway type.

**Table 2.** Healthcar euthiliztion before and after prosperity score matching.

| Pathway | Unmatched |  |  | Matched |  |  |
| --- | --- | --- | --- | --- | --- | --- |
|  | N of Encounters (mean per episode) | N of Labs (mean per episode) | N of Imaging (mean per 100 episode) | N of Encounters (mean per episode) | N of Labs (mean per episode) | N of Imaging (mean per 100 episode) |
| Dermatological symptoms |  |  |  |  |  |  |
| Digital | 1.98 | 0.90 | 1.38 | 2.00 | 0.96 | 1.53 |
| Traditional | 2.41 | 1.91 | 4.75 | 2.23 | 1.51 | 3.39 |
| Gastroenteritis |  |  |  |  |  |  |
| Digital | 1.85 | 0.86 | 0.62 | 1.78 | 0.98 | 0.64 |
| Traditional | 2.87 | 6.05 | 4.33 | 2.72 | 4.38 | 3.85 |
| Ophthalmological symptoms |  |  |  |  |  |  |
| Digital | 2.18 | 0.36 | 0.51 | 2.26 | 0.56 | 0.82 |
| Traditional | 2.49 | 1.56 | 3.91 | 2.46 | 1.17 | 3.70 |
| Respiratory infections |  |  |  |  |  |  |
| Digital | 2.34 | 1.00 | 2.24 | 2.34 | 1.00 | 2.24 |
| Traditional | 2.32 | 1.97 | 7.87 | 2.18 | 1.54 | 4.35 |
| Urinary tract infections |  |  |  |  |  |  |
| Digital | 2.59 | 4.72 | 1.04 | 2.62 | 4.90 | 1.16 |
| Traditional | 2.98 | 8.62 | 3.40 | 2.89 | 8.41 | 3.11 |

### Cost Com parison

Overall, episodes managed via the digital pathway incurred significantly lower costs compared to those in the traditional pathway, with a mean cost of €170.7 vs. €220.9, representing a 22.7% cost reduction (*P*<.001). This trend was consistently observed across the dataset (Table 3). On average, digital-first episodes were €50.17 less expensive per episode (*P*<.001) than those initiated through traditional care. These findings confirm the study hypothesis, demonstrating the cost-efficiency of digital pathways in primary care.

**Table 3.** Mean cost per episode by healthcare pathway (digital vs. traditional)

|  |  |  |  |  |
| --- | --- | --- | --- | --- |
| Category | Digital Pathway (€) | Traditional Pathway (€) | *P*-value | Cost Savings |
| Encounter costs, mean per episode | 166.54 | 212.68 | <.001 | 21.7% |
| Laboratory costs, mean per episode | 3.29 | 6.21 | <.001 | 47.0% |
| Imaging costs, mean per episode | 0.91 | 2.01 | <.001 | 54.7% |
| Total, mean per episode | 170.74 | 220.91 | <.001 | 22.7% |
| Note: Statistical comparisons performed using independent two-sample t-tests. Percent cost savings represent a relative reduction of digital compared to traditional care. |  |  |  |  |

Table 4 and Figure 2 details the cost outcomes by clinical presentation. In every category, the mean episode cost in the digital group was significantly lower than in the traditional group (*P*<.001 for each).

**Figure 1.**
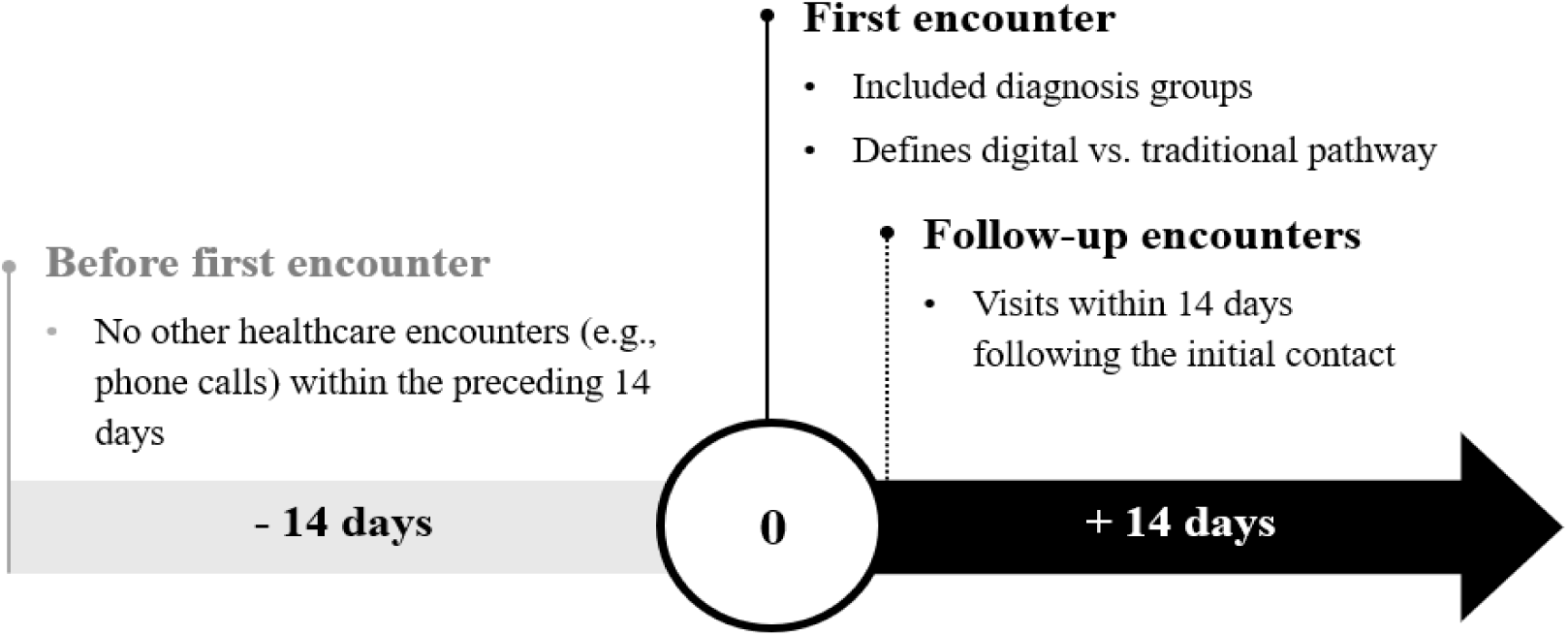
Conceptual definition of an episode of care.

**Figure 2.**
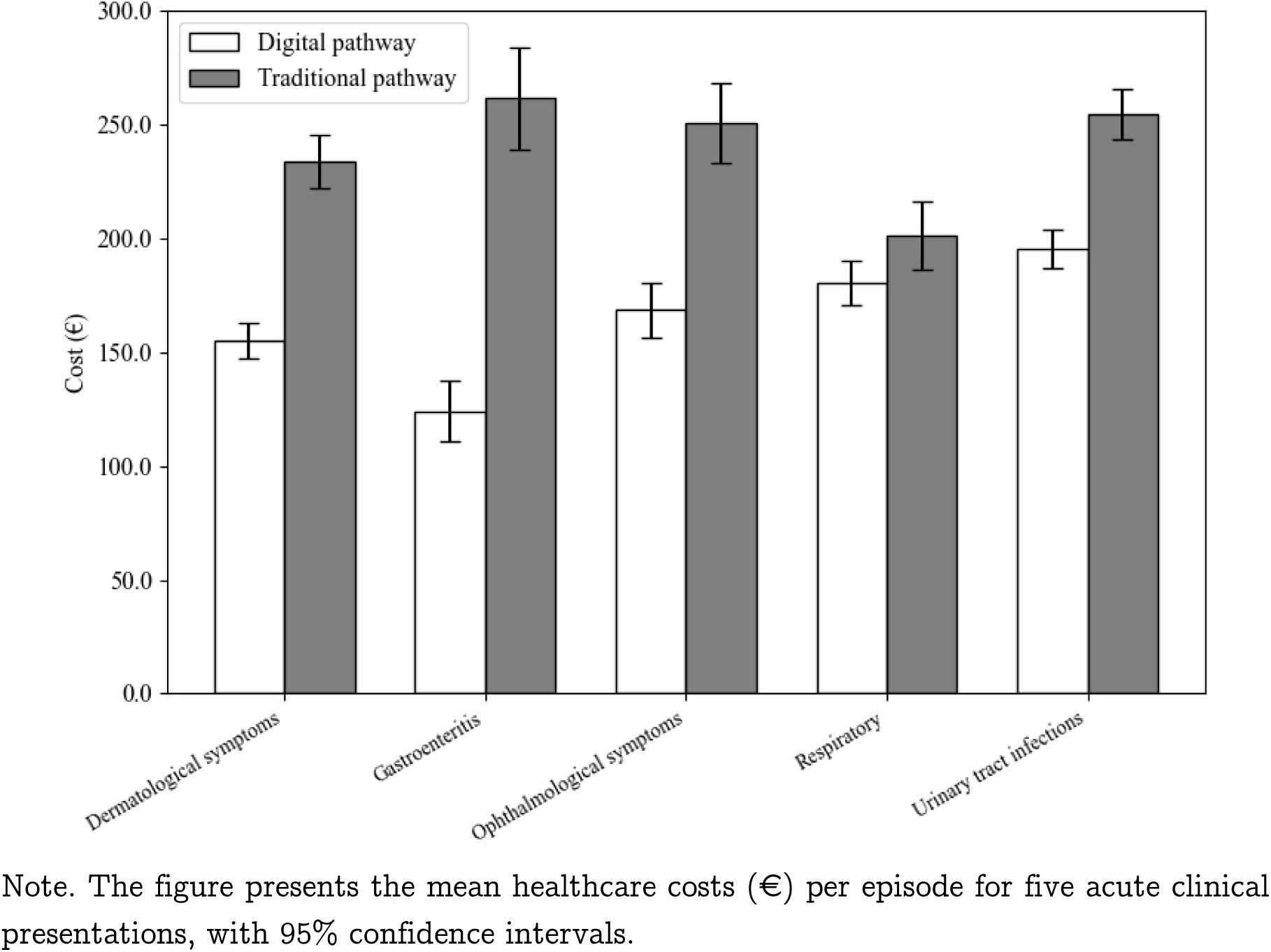
Mean total costs per episode by clinical presentation (digital vs. traditional pathway) 95% confidence intervals.

**Table 4.** Mean total cost per episode by clinical presentation and health care path way.

|  | Digital<br>Pathway,<br>mean per<br>episode (€) | Traditional<br>Pathway,<br>mean per<br>episode (€) | Cost<br>Savings | *t*-statistic | *P*-<br>value | N of<br>pairs |
| --- | --- | --- | --- | --- | --- | --- |
| Dermatological<br>symptoms | 155.21 | 233.81 | 33.6% | -29.72 | <.001 | 6 927 |
| Gastroenteritis | 124.02 | 261.36 | 52.5% | -10.34 | <.001 | 312 |
| Ophthalmological<br>symptoms | 168.38 | 250.61 | 32.8% | -13.40 | <.001 | 1 461 |
| Respiratory<br>infections | 180.27 | 200.96 | 10.3% | -9.20 | <.001 | 9 616 |
| Urinary tract<br>infections | 195.38 | 254.51 | 23.2% | -9.20 | <.001 | 1 381 |

Note: Each value represents the mean total cost per episode in Euros for that group and pathway. Percent reduction is calculated as (Trad – Digital)/Trad).

Across conditions, the gastroenteritis group showed the largest relative cost saving: digital gastroenteritis episodes cost about half as much as traditional episodes (∼52.5% less). This was followed by dermatologic symptoms with ∼33.6% savings and ophthalmologic symptoms with ∼32.8% savings. The urinary tract infections group saw about 23.2% lower costs with digital initiation. The respiratory infection group, which was the largest sample size, had a more modest 10.3% cost reduction via digital pathway.

In absolute terms, the cost differences were meaningful. For instance, an eye infection episode costs on average €82 less when handled digitally than traditionally. For skin issues (like eczema or rash management), digital episodes saved roughly €84 on average. These savings, multiplied over hundreds or thousands of cases, translate into substantial budget impact for the healthcare provider.

Looking at cost components, we found that the cost gap was primarily driven by differences in encounter costs. A digital consultation generally incurred a lower cost than an in-person visit, and digital users often resolved their issue with a single digital visit. In contrast, traditional pathway patients sometimes had an initial phone call followed by an in-person visit, effectively two encounters (and two charges). Laboratory and imaging utilization were relatively low in these acute conditions, but there was still a trend toward fewer tests in the digital group. For example, in urinary tract infections, clinicians in the traditional pathway more frequently order confirmatory lab tests and urine cultures. Overall, lab costs averaged slightly lower in digital episodes (€3.29 vs €6.21, a 47.0% saving). Imaging usage was minimal for these diagnoses, but even so, digital episodes had fewer X-rays (e.g. fewer sinus X-rays or chest X-rays for mild respiratory cases), with imaging costs roughly 50% lower (0.91€ vs 2.01€ on average). Notably, the digital pathway had lower utilization of laboratory tests and imaging for these acute conditions (as evidenced by lower associated costs), indicating a more streamlined care process.

### Sensitivity Analyses

The following sensitivity analyses were performed to assess the sensitivity of the results to different analytical choices: (a) varying the episode follow-up window to 7 and 30 (b) conducting an analysis on unmatched data utilizing multivariable regression; (c) adjusting cost assumptions to assess the influence of different pricing structures on the overall findings^27,28^; and (d) employing inverse probability of treatment weighting (IPTW) with truncated weights at the 1^st^ and 99^th^ percentile^29–31^. Sensitivity analyses all showed consistent cost reduction trends, with no analysis reversing the primary outcome, thereby supporting the robustness of the findings. See supplementary material B for details on sensitivity analyses.

## Discussion

Our study found that a digital-first primary care pathway reduced acute care episode costs by approximately 23% on average compared to traditional care, without increasing follow-up visits or unnecessary diagnostics. By analyzing over a year’s data from a Finnish primary care setting, we observed consistent cost reductions across multiple diagnostic categories. These findings reinforce prior research suggesting that digital health interventions can generate substantial economic benefits while maintaining clinical effectiveness^32^.

The digital pathway’s cost-effectiveness is influenced by several factors. One major factor is the reduction in overhead costs, as digital visits do not require physical room space, front-desk staff, or other infrastructure associated with in-person visits, which is reflected in the cost structure. In addition, the efficiency of encounters is evident; our data shows that digital consultations result in fewer interactions. Digital platforms effectively triage minor ailments that are self-limited, allowing them to be managed without an in-person exam. Digital clinicians can advise on supportive care and alert patients to red flags, therefore reducing unnecessary utilization. Moreover, conservative testing practices are encouraged in the digital pathway. Our results suggest that fewer laboratory and imaging studies are ordered in digital care settings. Providers may emphasize clinical history and adopt a watchful waiting approach for mild cases, whereas traditional clinical environments may have a tendency to perform additional diagnostic tests as a precaution. Overuse of low-value diagnostics in traditional settings can contribute to increased healthcare expenditures, whereas digital health platforms may inherently promote more judicious resource utilization, thereby mitigating these unnecessary diagnostic costs.

These findings contribute to the growing body of evidence on the effectiveness of digital consultations in primary care. Glock et al.^33^ demonstrated that eVisits in Swedish settings were suitable for managing uncomplicated conditions, with the majority of patients not requiring follow-up. Likewise, studies focusing on minor acute illnesses have generally reported no increase in downstream utilization following digital encounters^34–39^, with some reporting reduced service use^40^. However, earlier investigations have also noted increased follow-up visits and higher healthcare utilization following digital encounters^9,41–44^, particularly in cases of acute respiratory tract infections^45^. While few studies have assessed costs directly, those that have typically suggest favorable economic outcomes^5,6^. One exception is the study by Ashwood et al.^46^, which found higher costs for digital consultations in respiratory cases. Notably, respiratory infections represented the only clinical category in our study with slightly increased follow-up rates and the smallest relative cost savings.

The magnitude of cost savings in our study, ranging from 20% to 50% depending on condition, is higher than previously reported in some settings. For example, Buvik et al. observed a 19% cost reduction in orthopedic teleconsultations, contingent upon sufficient patient volume^47^. Gentili et al. found that over half of the studies in their review reported digital interventions as dominant—both more effective and less costly^5^. Our study provides concrete, real-world evidence of this dominance in a primary care context: the digital-first model achieved comparable clinical aims (resolution of minor acute illness) at lower cost.

On the other hand, a scoping review in Australia cautioned that telehealth’s system-wide savings were not always realized because of how services were funded and organized^48^. Importantly, our study setting enabled digital consultations to function as true substitutes rather than supplements to traditional care. The digital platform was fully integrated with clinical infrastructure, allowing for laboratory orders, imaging referrals, and escalation to in-person visits when necessary. This integration is critical—if digital services merely add another layer of care, overall utilization and costs may increase. In our context, where the same provider organization managed both digital and traditional services, care was coordinated, and duplication was avoided. Our data confirmed that digital encounters did not result in excess follow-ups or duplicated services, reinforcing the role of digital care as a substitute rather than a supplement.

These results have clear implications for healthcare policy. Scaling digital-first care models, particularly for conditions shown to be safely manageable online, could be a viable strategy for health systems seeking cost containment. With approximately 45,000 acute episodes per year in the study region, even partial uptake of the digital pathway could yield millions of euros in annual savings. Importantly, these savings do not require workforce reductions but could instead alleviate pressure to scale staffing proportionally with rising care demands— particularly as populations age and service needs increase. However, achieving these benefits at scale requires careful attention to clinical quality and health equity. Digital care protocols must include safeguards such as appropriate triage, red flag identification, and escalation procedures. Quality monitoring mechanisms are essential to ensure that cost reductions do not compromise clinical outcomes.

Another major implication is the need to address the digital divide. Our findings highlight a considerable age disparity in digital pathway utilization, with younger patients disproportionately using digital-first healthcare services. This aligns with prior evidence suggesting that older adults, individuals with limited internet access, and those with lower digital literacy are less likely to engage with digital health solutions ^8,9^. If these disparities remain unaddressed, the benefits of digital healthcare—such as improved access, convenience, and cost savings—may be inequitably distributed, reinforcing existing healthcare inequalities. However, digital health also has the potential to enhance accessibility, particularly in remote or underserved regions where traditional healthcare infrastructure is limited^49,50^. To ensure equitable adoption, targeted interventions should be implemented, including digital literacy programs tailored to older populations, user-friendly app designs that accommodate a wide range of technological proficiencies, and investments in infrastructure to expand internet connectivity and device accessibility in underserved areas.

By doing so, the cost benefits demonstrated here can be extended to a broad population without exacerbating inequity. In fact, successful integration of digital pathways could free up resources in traditional settings (e.g. less crowding in clinics), potentially improving access for those who do need or prefer in-person care (often older or sicker patients). This synergy can enhance overall system efficiency and patient satisfaction.

Our results also feed into the broader discussion of healthcare system reform. Finland’s healthcare reform seeks cost savings to rein in deficits, and digital health has been highlighted as a key strategy. The evidence from this study supports those strategic directions – a digital-first primary care model can indeed contribute to cost containment. Policymakers and administrators can leverage these findings to justify investments in digital platforms. While there is an upfront cost to building and maintaining apps and IT infrastructure, the return on investment appears favorable given the per-episode savings. Additionally, while our analysis focused on the provider perspective, societal benefits such as reduced patient travel, lower absenteeism, patient preference^51–54^ and environmental advantages^55,56^ further strengthen the case for digital pathways, even though these were not quantified in this study.

Furthermore, ensuring that digital consultations hold equivalent status to phone consultations in clinical workflows is crucial to reducing unnecessary follow-up contacts and optimizing resource use. Our findings support the adoption of an omnichannel care model, where patients can receive effective, cost-efficient care through multiple modalities, allowing for greater flexibility while maintaining high-quality service delivery.

### Limitations

This study has several limitations to acknowledge. First, although we used propensity score matching to balance observed characteristics between digital and traditional pathway users, residual confounding may remain. Unmeasured factors—such as clinical complexity, digital literacy, or patient preference—may have influenced both care pathway selection and healthcare-seeking behavior. While we matched prior utilization (a proxy for healthcare engagement), these unmeasured differences could still impact outcomes. Nonetheless, the observed cost differences were substantial, making it unlikely that residual confounding fully explains the findings.

Second, as a cost-minimization analysis, we did not directly assess clinical outcomes such as symptom resolution, patient satisfaction, or complication rates. We assumed equivalent outcomes between pathways based on clinical judgment and similar follow-up rates. While prior studies suggest that telemedicine provides comparable outcomes for acute conditions, subtle differences in quality or patient experience cannot be ruled out.

Third, the study is region-specific, focusing on Päijät-Häme in Finland, within a healthcare system that integrates digital services into public primary care. While the findings are likely generalizable to settings with similar infrastructure and care models, they may not fully apply to healthcare systems with different payment structures, such as purely fee-for-service models. Differences in reimbursement incentives, physician practice patterns, or digital adoption rates could lead to variations in cost-effectiveness across different healthcare environments.

Fourth, the time horizon was relatively short (14 days, with a 30-day sensitivity analysis), capturing only the acute episode costs. While this timeframe is appropriate for evaluating short-term healthcare expenditures, it does not account for any potential long-term cost implications. If digital consultations led to increased prescribing patterns or downstream healthcare utilization, such effects would not be captured in this analysis. However, our data did not indicate concerning trends in follow-up care rates, suggesting that cost savings were not achieved at the expense of additional downstream resource use. Longer-term studies are warranted, particularly to assess the economic impact of digital care pathways for chronic disease management.

Fifth, cost estimates are inherently subject to variability. Our analysis is based on publicly available regional cost data, which represents average production costs rather than real-time, transaction-level expenditures. While this approach aligns with economic evaluation best practices, healthcare cost structures vary across systems, and the results may not fully capture hidden or indirect costs associated with digital and traditional pathways.

Sixth, episode-based costing may include services unrelated to the acute condition. However, this applies equally to both groups and is unlikely to introduce systematic bias.

Seventh, the study examined costs solely from the healthcare provider’s perspective, excluding societal costs such as patient travel expenses, lost work time, or productivity losses. Given that digital consultations eliminate the need for physical travel and reduce time burdens for patients, incorporating these factors in future cost-benefit analyses would likely further strengthen the case for digital-first pathways.

## Conclusion

This study supports the view that digital-first primary care pathways can significantly reduce healthcare costs for acute conditions without a significant increase in follow-up visits. In the examined Finnish health system, digital care pathways lowered the cost of managing common infections and minor illnesses by approximately 20–50%, depending on the condition. These findings support the integration of digital health solutions as a scalable strategy for improving healthcare system efficiency.

Beyond the advantages of convenience and accessibility, this study highlights the economic rationale for expanding digital-first healthcare models. Healthcare leaders and policymakers should recognize that digital health is not only a means of enhancing patient access but also a viable approach to cost containment. However, achieving widespread adoption while ensuring equitable access and maintaining clinical quality will be critical.

With thoughtful implementation, digital pathways can contribute to slowing the rise in healthcare expenditures—a key priority for the long-term sustainability of health systems in Finland and globally.

## Data Availability

All data produced during this study are included in this published article or supplementary attachments. Due to privacy regulations, patient-level data cannot be shared.

## Acknowledgments

The authors acknowledge the support of Mehiläinen Oy, Harjun Terveys oy, and Päijät-Häme Wellbeing Services County. The analysis was conducted as part of the lead author’s master’s thesis at Hanken School of Economics. The authors used ChatGPT (OpenAI)^57^ to enhance the clarity and coherence of the manuscript; however, all content was critically reviewed, revised, and approved by the authors, who assume full responsibility for the accuracy and integrity of the final publication.

## Code Availability

The underlying code for this study is available upon request.

## Authors’ Contributions

A.D. led the conceptualization and design of the study, conducted the data analysis, and drafted the initial manuscript. S.J. supervised the research process, provided methodological guidance, and critically revised the manuscript. T.K. contributed to the study’s digital healthcare framework, advising on data structuring, and contributed to manuscript development. P.O. provided strategic input on study design, facilitated access to relevant data sources, and contributed to manuscript development. All authors contributed to the interpretation of findings and approved the final version for submission.

## Declaration of Interest

The authors declare the following competing interests: All authors are employed by Mehiläinen, the healthcare provider that developed and operates the digital clinic evaluated in this study. Some authors hold personal financial interests in Mehiläinen. The study was funded by Mehiläinen, and the leadership of Harjun terveys provided institutional support throughout the research process.

## Ethical Approval

This study was based on pseudonymized administrative healthcare data and did not involve direct interaction with patients. As such, individual patient consent or ethical approval was not required due to the Finnish Secondary Use act of personal data. The study was approved by the regional healthcare authority, and data use agreements were in place. The research complies with relevant data protection regulations, including the Finnish *Act on the Secondary Use of Health and Social Data (552/2019)* and the *EU General Data Protection Regulation (GDPR 2016/679)*. Research permission was obtained from Päijät-Häme Wellbeing Services County in November 2024, following the submission of a detailed research proposal and supporting applications.

## Supplementary M aterial A

**Supplementary Table A1.** Diagnosis codes included in the study.

Supplementary Material A
| Supplementary Table A1. Diagnosis codes included in the study. |  |  |
| --- | --- | --- |
| Diagnosis codes | ICD-10 | ICPC-2 |
| Dermatological symptoms | L | S01-S76, S81-S99 |
| Gastroenteritis | A08-A09 | D73 |
| Ophthalmological symptoms | H00, H01, H04.10, H10, H11.0, H11.3 | F01, F02, F03, F13, F15, F16, F29, F70, F71, F72, F73, F75 |
| Respiratory infections | J00-J22 | R05, R08, R09, R21, R25, R71, R72, R74, R75, R76, R77, R78, R80, R83 |
| Urinary tract infections | N30.0, N30.9 | U01, U02, U05, U07, U13, U70, U71, U72 |
| Note. The selected diagnoses represent the most frequently encountered acute conditions managed through the digital pathway, comprising approximately 50% of all encounters within this care model. These conditions were chosen based on their high prevalence, suitability for remote assessment, and clinical relevance for both digital and traditional care pathways. The inclusion of both ICD-10 and ICPC-2 codes ensures comprehensive classification and facilitates comparability across different healthcare coding systems. |  |  |

**Supplementary Table A2.**

| Supplementary Table A2. |  |
| --- | --- |
|  | Päijät-Häme |
| Nurses' digital appointment | €61 |
| Nurses' appointment | €98 |
| Doctors' consultation | €55 |
| Doctors' digital appointment | €100 |
| Doctors' physical appointment | €166 |
| Note. Confirmed by the Wellbeing Services County. |  |

**Supplementary Table A3.**
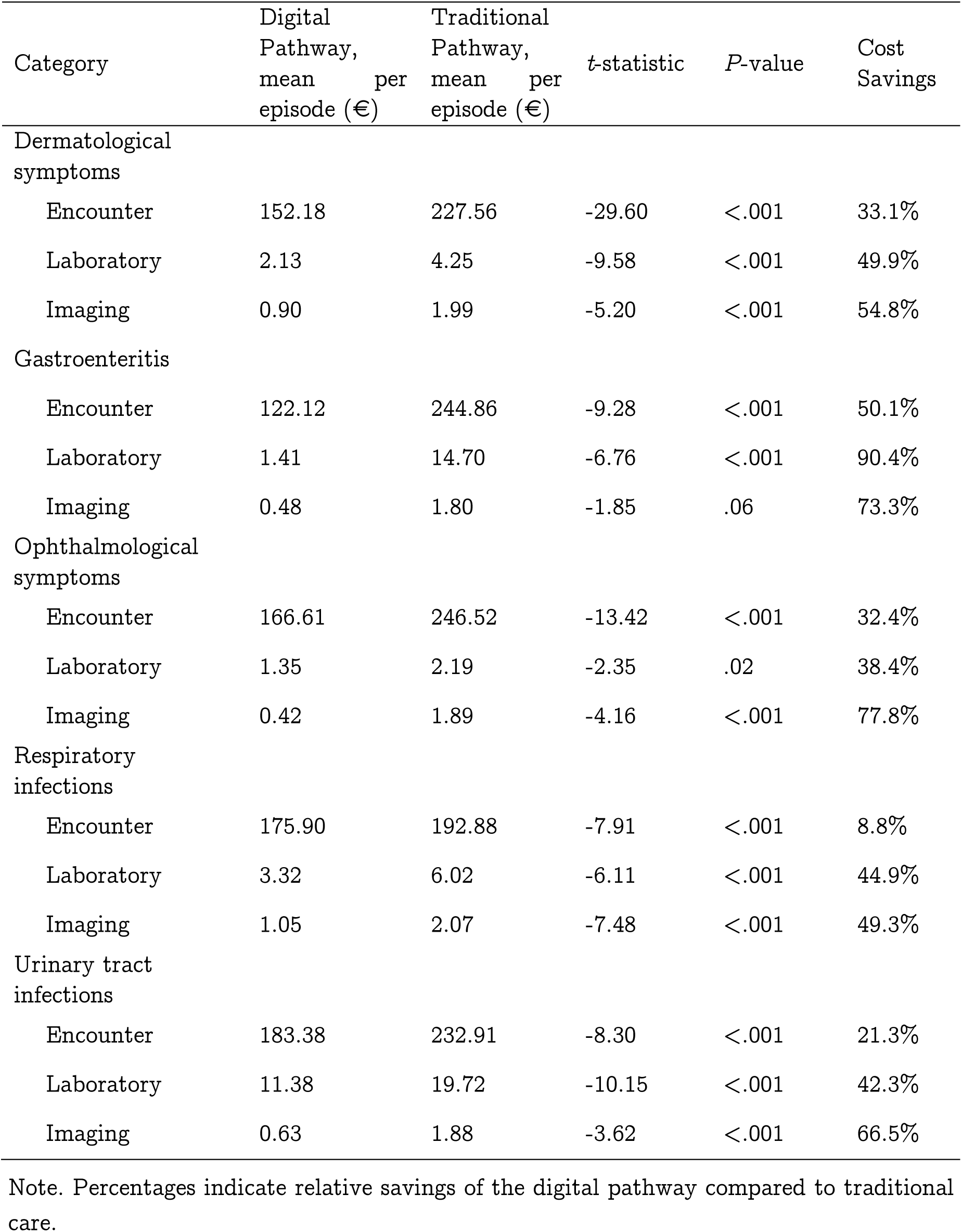
Breakdown of mean episode costs (€) by clinical presentation and healthcare pathway (digital vs. traditional)

## Supplementary M aterial B

**Supplementary Table B1.** Sensitivity analysis: CMA results using unmatched patient groups.

Supplementary Material B
|  |  |  |  |  |  |
| --- | --- | --- | --- | --- | --- |
|  | Digital Pathway, total costs (mean per episode, €) | Traditional Pathway, total costs (mean per episode, €) | *t*-statistic | *P*-value | Cost Savings |
| Dermatological symptoms | 152.72 | 254.21 | -48.30 | <.001 | 39.9% |
| Gastroenteritis | 130.95 | 283.17 | -12.58 | <.001 | 53.8% |
| Ophthalmological symptoms | 158.92 | 257.19 | -22.11 | <.001 | 38.2% |
| Respiratory infections | 180.27 | 218.56 | -18.87 | <.001 | 17.5% |
| Urinary tract infections | 191.37 | 260.32 | -12.27 | <.001 | 26.1% |
| Note: This table presents the results of a sensitivity analysis comparing mean total episode costs (€) between digital and traditional healthcare pathways without propensity score matching (unmatched analysis). |  |  |  |  |  |

**Supplementary Table B2.** Sensitivity analysis: CMA results using same patient Digital.

| Supplementary Table B2. Sensitivity analysis: CMA results using same patient |  |  |  |  |  |  |
| --- | --- | --- | --- | --- | --- | --- |
|  | Digital Pathway, mean total cost per episode (€) | Traditional Pathway, mean total cost per episode (€) | N of pairs | *t*-statistic | *P*-value | Cost Savings |
| Dermatological symptoms | 237.94 | 329.50 | 1 223 | -7.17 | <.001 | 27.8% |
| Gastroenteritis | 216.94 | 295.87 | 13 | -1.12 | .28 | 26.7% |
| Ophthalmological symptoms | 212.13 | 278.90 | 116 | -3.19 | .002 | 23.9% |
| Respiratory infections | 291.68 | 309.97 | 1 355 | -1.86 | .06 | 5.9% |
| Urinary tract infections | 288.54 | 325.65 | 136 | -1.13 | .26 | 11.6% |
| Note: This sensitivity analysis compares costs within the same patients who used both digital and traditional healthcare pathways, controlling patient-level variability. |  |  |  |  |  |  |

**Supplementary Table B3.** Sensitivity analysis: CMA results using alternative unit costs (THL pricing catalog)

|  |  |  |  |  |  |  |
| --- | --- | --- | --- | --- | --- | --- |
|  | Digital<br>Pathway,<br>mean total<br>cost per<br>episode (€) | Traditional<br>Pathway,<br>mean total<br>cost per<br>episode (€) | *t*-statistic | *P*-value | N of<br>Pairs | Cost<br>Savings |
| Dermatological symptoms | 80.15 | 127.13 | -31.85 | <.001 | 6 927 | 37.0% |
| Gastroenteritis | 58.01 | 136.47 | -10.86 | <.001 | 312 | 57.5% |
| Ophthalmological symptoms | 99.18 | 142.61 | -12.37 | <.001 | 1 461 | 30.5% |
| Respiratory infections | 87.82 | 106.11 | -14.49 | <.001 | 9 616 | 17.2% |
| Urinary tract infections | 108.06 | 139.54 | -8.71 | <.001 | 1 381 | 22.6% |
| Note: Inflation-adjusted standardized national pricing data from the Finnish Institute for Health and Welfare (THL). |  |  |  |  |  |  |

**Supplementary Table B4.** Sensitivity analysis: CMA results for 7-day episode window Digital.

| Supplementary Table B4. Sensitivity analysis: CMA results for 7-day episode window |  |  |  |  |  |  |
| --- | --- | --- | --- | --- | --- | --- |
|  | Digital<br>Pathway,<br>mean total<br>cost per<br>episode (€) | Traditional<br>Pathway,<br>mean total<br>cost per<br>episode (€) | *t*-statistic | *P*-value | N of<br>Pairs | Cost<br>Savings |
| Dermatological symptoms | 122.27 | 204.36 | -41.78 | <.001 | 7 502 | 40.2% |
| Gastroenteritis | 102.23 | 229.00 | -12.29 | <.001 | 355 | 55.4% |
| Ophthalmological symptoms | 142.19 | 231.55 | -19.59 | <.001 | 1 766 | 38.6% |
| Respiratory infections | 145.79 | 187.78 | -22.43 | <.001 | 9 321 | 22.4% |
| Urinary tract infections | 162.63 | 225.67 | -12.71 | <.001 | 1 516 | 27.9% |

**Supplementary Table B5.** Sensitivity analysis: CMA results for 30-day episode window Digital.

| Supplementary Table B5. Sensitivity analysis: CMA results for 30-day episode window |  |  |  |  |  |  |
| --- | --- | --- | --- | --- | --- | --- |
|  | Digital<br>Pathway,<br>mean total<br>cost per<br>episode (€) | Traditional<br>Pathway,<br>mean total<br>cost per<br>episode (€) | *t*-<br>statistic | *P*-value | N of<br>Pairs | Cost<br>Savings |
| Dermatological symptoms | 180.68 | 285.67 | -28.69 | <.001 | 6 058 | 36.8% |
| Gastroenteritis | 151.13 | 295.49 | -9.23 | <.001 | 299 | 48.9% |
| Ophthalmological symptoms | 183.07 | 287.02 | -15.16 | <.001 | 1 562 | 36.2% |
| Respiratory infections | 194.42 | 242.10 | -15.66 | <.001 | 7 506 | 19.7% |
| Urinary tract infections | 224.54 | 303.38 | -9.69 | <.001 | 1 297 | 26.0% |

**Supplementary Table B6.** Sensitivity analysis: Mean episode costs using inverse probability of treatment weighting (IPTW)

|  |  |
| --- | --- |
|  | Weighted<br>Mean Cost (€) |
| Dermatological symptoms |  |
| Digital | 169.97 |
| Traditional | 248.17 |
| Gastroenteritis |  |
| Digital | 133.93 |
| Traditional | 267.07 |
| Ophthalmological symptoms |  |
| Digital | 166.01 |
| Traditional | 245.53 |
| Respiratory infections |  |
| Digi | 189.59 |
| Traditional | 212.49 |
| Urinary tract infections |  |
| Digital | 194.61 |
| Traditional | 256.03 |

Note: Weighted mean total costs per episode calculated via inverse probability of treatment weighting (IPTW), an approach estimating the average treatment effect across the full study population. Weights were truncated at the 1st and 99th percentiles to mitigate extreme values.

**Supplementary Table B7.**
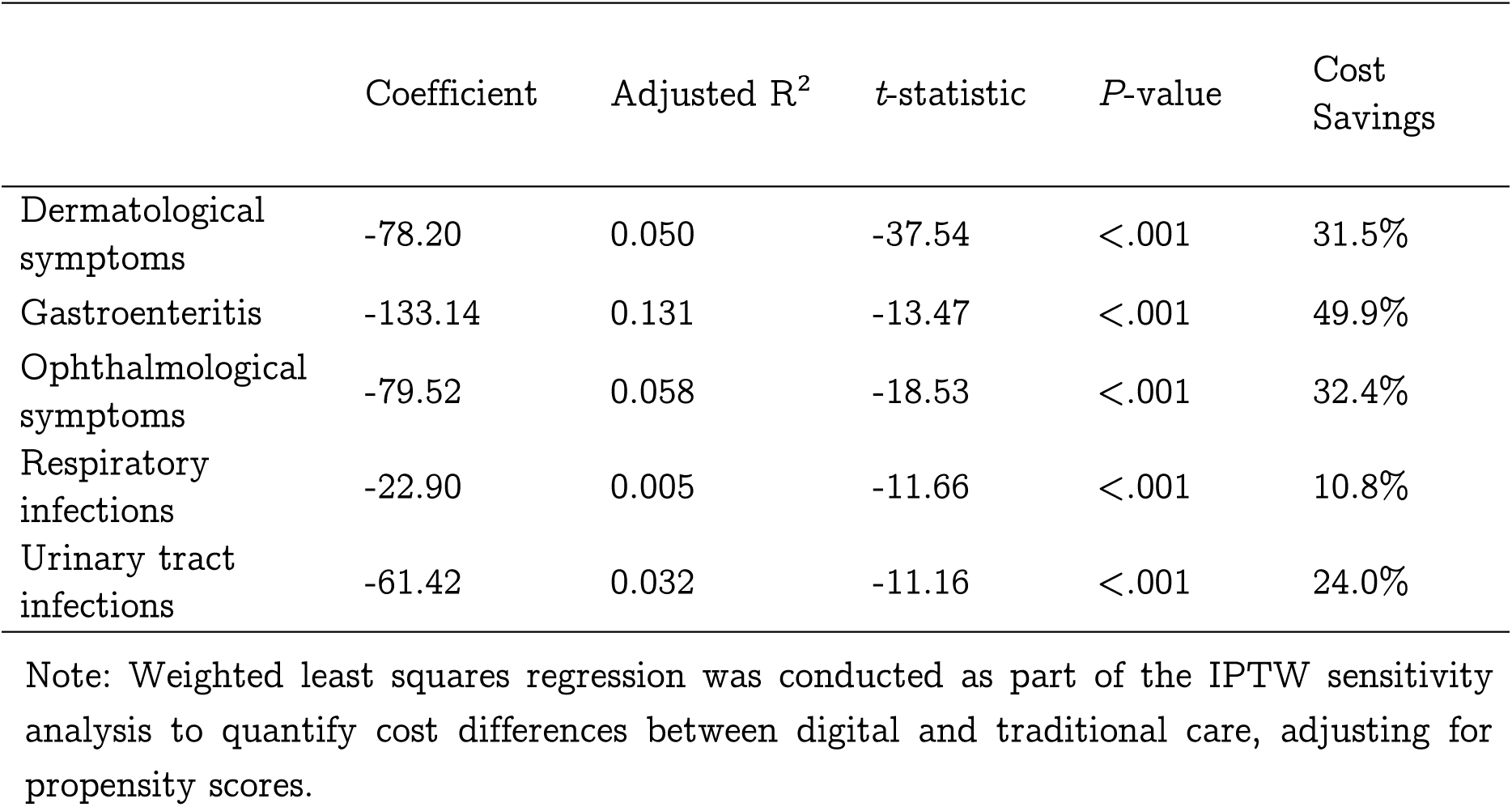
Sensitivity analysis: Weighted Least Squares (WLS) regression results (IPTW analysis)

|  |  |  |  |  |  |
| --- | --- | --- | --- | --- | --- |
|  | Coefficient | Adjusted R<sup>2</sup> | *t*-statistic | *P*-value | Cost Savings |
| Dermatological symptoms | -78.20 | 0.050 | -37.54 | <.001 | 31.5% |
| Gastroenteritis | -133.14 | 0.131 | -13.47 | <.001 | 49.9% |
| Ophthalmological symptoms | -79.52 | 0.058 | -18.53 | <.001 | 32.4% |
| Respiratory infections | -22.90 | 0.005 | -11.66 | <.001 | 10.8% |
| Urinary tract infections | -61.42 | 0.032 | -11.16 | <.001 | 24.0% |
| Note: Weighted least squares regression was conducted as part of the IPTW sensitivity analysis to quantify cost differences between digital and traditional care, adjusting for propensity scores. |  |  |  |  |  |

## Notes

### Funding Statement

This study was funded by the service provider in question.

### Author Declarations

Päijät Häme Wellbeing Services County Research Permit, Case Number HA/85/07.01.04.05/2024 Ethical approval waived; retrospective registry-based study.

